# Association of natural induced antibodies against select Group B streptococcus surface proteins and invasive disease in early infancy

**DOI:** 10.1101/2024.07.19.24310719

**Authors:** Sonwabile Dzanibe, Alane Izu, Ziyaad Dangor, Gaurav Kwatra, Peter V. Adrian, Sheila Z. Kimaro Mlacha, Shabir A. Madhi

## Abstract

**Background:** Group B *streptococcus* (GBS) surface protein epitopes are potential targets for development of vaccines that could confer protection against invasive GBS disease (IGbsD) irrespective of the capsular serotype. The aim of this study was to determine the association of natural acquired mother-newborn GBS surface proteins specific serum IgG and IGbsD during early infancy (<90 days age).

**Methods:** Clinical GBS isolates from 81 women who delivered either infant with IGbsD (n=38) and healthy controls (n=43) were assessed for surface expression of proteins Sip, GBS0393 and GBS206 using flow cytometry. Serum IgG titres to Sip, GBS0393 and GBS206 surface proteins were measured in paired maternal-infant sera on multiplex Luminex platform to determine IgG titres associated with reduce risk of IGbsD caused by GBS strains expressing homotypic protein.

**Results:** Infant sera IgG GMT in IGbsD cases caused by strains expressing Sip and GBS2016 were lower (64 and 61 U/ml, respectively) compared to healthy controls born to women colonized by strains expressing the respective proteins (145 and 151 U/mL, respectively, p<0.01). Moreover, increasing infant antibody titres against Sip and GBS2016 were associated with ≥80% adjusted disease risk (ADR) reduction to GBS isolates expressing homotypic proteins. Among women colonized with GBS isolates expressing GBS2106, mothers of cases had lower GBS2016-specific IgG GMT (249 U/mL) compared to mothers of controls (163 U/mL, adj-p=0.023).

**Conclusion:** Increasing infant IgG titres to GBS2106 and Sip was associated with reduced IGbsD risk and therefore warranting further investigations as potential GBS vaccines and/or protein conjugants.

## 1. Introduction

Group B *Streptococcus* (GBS) is a leading cause of neonatal invasive disease, with a case fatality risk between 2-20% (1–4). Invasive GBS disease (IGbsD) is traditionally classified as early-onset (EOD, <7 days of age) or late-onset disease (LOD, 7-90 days of age), reflecting possible differences in acquisition of GBS by the neonate and pathogenesis of illness. Maternal vaccination against GBS could theoretically prevent IGbsD in their infants, as suggested by reports on the inverse association between serotype-specific capsular antibody and risk for invasive GBS disease in infants born to women colonised with the homotypic serotype. (5–10). GBS capsular polysaccharide conjugated to protein carrier are being developed as vaccine candidates, including an experimental multivalent vaccine that was evaluated for safety and immunogenicity in pregnant and non-pregnant women. (11–14). The polysaccharide-protein conjugate vaccines are, however, serotype specific (currently targeting six of the 10 known GBS serotypes) (14,15), and have been expensive to produce.

Theoretically GBS common protein antigens either alone, or as protein conjugates to the capsular polysaccharide, could expand protection against a greater diversity of serotypes. Also, the protein epitopes could elicit antibody responses that may act synergistically with serotype-specific capsular antibody, enhancing the efficacy compared to conjugate vaccines using non-GBS protein epitopes as the conjugating protein.

Here, we investigated the absolute disease risk reduction (ADRR) of paired infant-maternal serum IgG to GBS surface proteins Sip, GBS0393 and GBS2106 between IGbsD due to strains expressing homotypic protein antigens in young infants (<90 days age) and matched controls. We show that increasing antibody titres against a novel GBS2106 surface protein in infant sera was associated with ≥80% IGbsD risk reduction.

## 2. Methods

### 2.1. Study population

We retrospectively analysed serum samples from young infants (<90 days age) and their mothers (Table 1) that were previously enrolled in a case-control study investigating the association of capsular serotype-specific antibody and invasive GBS disease (10). Briefly, infants <90 days of age with GBS cultured from either blood, cerebrospinal fluid or other normally sterile sites were enrolled together with their mothers. Healthy young infants without invasive GBS disease were enrolled as the control group, matched for maternal HIV status, maternal age (±2.5 years), gestational age (≥37 weeks or ± 2 weeks for preterm), and age of disease onset in the cases (<6 days for EOD or ±14 days for LOD). Lower vaginal and rectal swabs were collected form the mothers at enrolment to determine GBS colonisation status and serotyping of GBS isolates was done by latex agglutination test as described (10). We limited out analysis to 83 women who had term deliveries (≥37 weeks gestation) and were matched for colonizing serotype between mothers of cases and controls prior assessing protein surface expression.

**Table 1.** Demographic characteristics for mother-infant pairs colonised with group B Streptococcus isolates expressing either Sip, gbs2106 or gbs0393 on the bacterial surface.

|  | **SIP** |  |  | **GBS2106** |  |  | **GBS0393** |  |  |
| --- | --- | --- | --- | --- | --- | --- | --- | --- | --- |
|  | Case<br>(n=27) | Colonized<br>Control<br>(n=30) | p-value | Case<br>(n=38) | Colonized<br>Control<br>(n=38) | p-value | Case<br>(n=34) | Colonized<br>Control<br>(n=37) | p-value |
| **Maternal** |  |  |  |  |  |  |  |  |  |
| Median age in years<br>(IQR) | 25.8<br>(22.5-31.9) | 25<br>(21.1-30.1) | 0.313 | 28<br>(23.5-31.6) | 25.3<br>(21.1-29.4) | 0.106 | 27.2<br>(23.5-31.6) | 25.9<br>(21.4-30.4) | 0.400 |
| Median parity (IQR) | 1 (0-2) | 0 (0-1) | 0.163 | 1 (0-2) | 0 (0-1) | 0.028 | 1 (0-2) | 0 (0-1) | 0.056 |
| Median gravidity (IQR) | 2 (1-3) | 1.5 (1-2) | 0.145 | 2 (1-3) | 2 (1-2) | 0.048 | 2 (1-3) | 2 (1-2) | 0.086 |
| PROM≥18h (%) | 3 (13.6) | 1 (3.6) | 0.308 | 3 (9.7) n=31 | 2 (5.9) n=34 | 0.663 | 3 (10.0) n=30 | 2 (6.1) n=33 | 0.662 |
| Colonising serotype (%) |  |  |  |  |  |  |  |  |  |
| Ia | 11 (40.7) | 15 (39.5)<sup>†</sup> |  | 14 (36.8) | 16 (34.8)<sup>†</sup> |  | 14 (41.2) | 19 (42.2)<sup>†</sup> |  |
| Ib | 2 (7.4) | 2 (5.3)<sup>†</sup> |  | 2 (5.3) | 3 (6.5)<sup>†</sup> |  | 2 (5.9) | 3 (6.7)<sup>†</sup> |  |
| II | 1 (3.7) | 2 (5.3)<sup>†</sup> |  | 1 (2.6) | 3 (6.5)<sup>†</sup> |  | 1 (2.9) | 3 (6.7)<sup>†</sup> |  |
| III | 10 (37.0) | 12 (31.6)<sup>†</sup> |  | 18 (47.4) | 17 (37.0)<sup>†</sup> |  | 14 (41.2) | 13 (28.9)<sup>†</sup> |  |
| V | 3 (11.1) | 7 (18.4)<sup>†</sup> | 0.932 | 3 (7.9) | 7 (15.2)<sup>†</sup> | 0.732 | 3 (8.8) | 7 (15.6)<sup>†</sup> | 0.747 |
| HIV-infected (%) | 11 (40.7) | 10 (33.3) | 0.594 | 18 (47.4) | 12 (31.6) | 0.240 | 17 (50.0) | 12 (32.4) | 0.154 |
| *Median CD4 cells/μL (IQR)* | 387<br>(260-659) | 550<br>(339-680) | 0.518 | 385<br>(294-605) | 384<br>(308-685) | >0.999 | 383<br>(280-620) | 381<br>(295-515) | 0.796 |
| **Infant** |  |  |  |  |  |  |  |  |  |
| *Median gestational age (IQR)* | 40<br>(38.5-40.5) | 39.2<br>(38.5-40.5) | 0.749 | 40<br>(39-40.5) | 40<br>(39-40.4) | 0.527 | 40<br>(38.7-40.6) | 39.5<br>(39-40.3) | 0.544 |
| *37 weeks (%)* | 0 (0.0) | 0 (0.0) | >0.999 | 0 (0.0) | 0 (0.0) | >0.999 | 0 (0.0) | 0 (0.0) | >0.999 |
| *Median birth weight in grams (IQR)* | 3010<br>(2800-3243) | 3080<br>(2874-3446) | 0.392 | 3080<br>(2846-3365) | 3110<br>(2918-3484) | 0.557 | 3060<br>(2809-3388) | 3105<br>(2925-3450) | 0.530 |
| *<2500 (%)* | 1 (3.7) | 0 (0.0) | 0.474 | 1 (2.6) | 0 (0.0) | >0.999 | 1 (2.9) | 0 (0.0) | 0.479 |
| *Invasive disease onset* |  |  |  |  |  |  |  |  |  |
| *LOD (%)* | 12 (44.4) | 15 (50.0) |  | 18 (47.4) | 19 (50.0) |  | 15 (44.1) | 17 (45.9) |  |
| *EOD (%)* | 15 (55.6) | 15 (50.0) | 0.792 | 20 (52.6) | 19 (50.0) | >0.999 | 19 (55.9) | 20 (54.1) | >0.999 |

‡There were four women colonised with multiple GBS serotypes: Ia and Ib; Ia and V; II and III; II and V.
\*There was no difference between expression of protein and disease status, stratified by serotype

### 2.2. Recombinant proteins

Putatively immunogenic surface proteins were identified *in silico* using the genome sequence of *Streptococcus agalactiae* NEM316 as previously described (16). Genes encoding proteins Sip, GBS0393, GBS1356 and GBS2106 were cloned into *E. coli*, recombinantly produced and purified using Ni^2+^ affinity columns for serological testing as previously described (16).

### 2.3. Quantification of antibodies against GBS surface proteins

Purified recombinant proteins were desalted using PD-10 columns (GE Healthcare, Germany) and 50 µg/mL of each protein was coupled to unique microbeads (Bio-Rad, Hercules, CA) using an adaptation of Pickering et al. protocol (17). Sera was diluted 1:200 in PBS, 10% fetal bovine serum, 0.05% NaN_3_ (PBS-FN) and mixed with an equal volume of protein-coupled beads. Bound antibodies were detected with goat anti-human IgG conjugated to R-phycoerythrin (Jackson ImmunoResearch, USA). Protein specific IgG in maternal and infant sera were measured using Bio-plex 200 (Bio-Rad, Hercules, CA) in a multiplex immunological assay as described (16), and titres reported as arbitrary units per mL (U/mL).

### 2.4. Generation of GBS protein specific polyclonal antibodies

Purified GBS proteins Sip, GBS0393, GBS1356 and GBS2106 were used to generate protein specific polyclonal antibodies in 6-week old female New Zealand white rabbits. The rabbits were injected subcutaneously with 250 µg of the recombinant proteins emulsified in complete Freud’s adjuvant followed by two booster doses containing incomplete Freud’s adjuvant at day 28 and 42 post primary immunisation. Blood was harvested by exsanguination at day-56 and isolated sera stored at −20°C.

### 2.5. Bacterial surface staining

Clinical GBS isolates were cultured at 37°C in Todd Hewitt medium (Davies Diagnostics, South Africa) until OD_600nm_ of 0.5. The cultures were washed with PBS and fixed using PBS, 0.1% paraformaldehyde solution at 37°C for 1 h and at 4°C for ∼16 h. The fixed bacterial cells were washed twice with PBS and 50 µL of the bacterial cells (OD_600nm_ of 0.1) were incubated with 50 µL of PBS, 20% fetal calf serum and 0.1% bovine serum albumin (PBS-FB) for 20 min. Bacterial samples were mixed with 100 µL of rabbit sera diluted 1:200 in PBS-FB and incubated for 1 h at 4°C. Cells were washed with PBS, 0.1% bovine serum albumin and stained with 50 µL of anti-rabbit IgG conjugated to R-phycoerythrin (Jackson ImmunoResearch, USA) diluted 1:200 in PBS-FB. Stained bacterial cells were washed, resuspended in 200 µL and inspected by flow cytometry (FACS Calibur, Becton-Dickinson Bioscience). Results were analyzed using FlowJo software (version 10.1r5, Tree Star Inc., CA).

### 2.6. Statistical analysis

The data was analysed using R (version 3.5 R Core Team, Vienna, Austria) and STATA software (version 13.0 Stata-Corp, Tx USA). IgG antibody titres measured in both infant and maternal sera were log10 transformed for parametric statistical comparisons and reported as geometric mean titres (GMT, U/mL). Student’s t-test was used to compare serum IgG GMT in infants without IGbsD born to colonized (colonized controls) and in infants with IGbsD (cases); and similarly, so in their mothers. All IgG titres to Sip, GBS0393 and GBS2106 were compared between cases and controls colonised with GBS strains expressing homotypic protein. Maternal age, maternal HIV status, gestational age and premature rupture of membrane were adjusted using multivariate regression. Fisher’s exact test was used to compare proportions between groups. P-value < 0.05 was considered statistically significant.

Reverse cumulative plots (RCP) and stratified log rank tests were used to compare IgG distribution between cases and controls colonised with GBS strains expressing homotypic proteins. Where significant differences were found, we estimated the Absolute Disease Risk Reduction (ADRR) over a range of possible antibody concentrations using both a parametric (18) and non-parametric approach (19) For parametric estimation, we fit a Bayesian model assuming antibody concentrations for both cases and controls follow a Weibull distribution and disease incidence follows a Beta(25,25000) distribution, which is centred at 0.001.

### 2.7. Ethics

Informed consent was obtained from all study participants at enrolment in the original study, which included consent for future use of archived samples for further studies contingent upon approval by the Ethics committee. This study was approved by the Human Research Ethics Committee at the University of Witwatersrand (Approval number: M140902). Animal use for generating polyclonal antibodies was approved by the Animal Ethics Screening Committee of the University of Witwatersrand (Approval number: 2015/05/20/B).

## 3. Results

### Surface protein expression on clinical GBS isolates

Using flow cytometry, we assessed the expression of Sip, GBS0393, GBS1356 and GBS2106 on the surface of *S. agalactiae* NEM316, a strain from which the recombinant proteins were cloned. Bacteria were stained with rabbit anti-sera collected pre- and post-immunisation, and the difference in fluorescence signal intensity used to measure accessibility of protein on the surface of GBS. Antibody binding on the surface of GBS bacteria were ≥6-fold higher in antigen specific sera compared to pre-immunisation sera (control) for Sip, GBS0393 and GBS2106 (Figure 1A). No difference in fluorescence intensity was observed between control (pre-immunization) and anti-sera (post-immunization) for GBS1356 and therefore omitted from further analysis.

**Figure 1.**
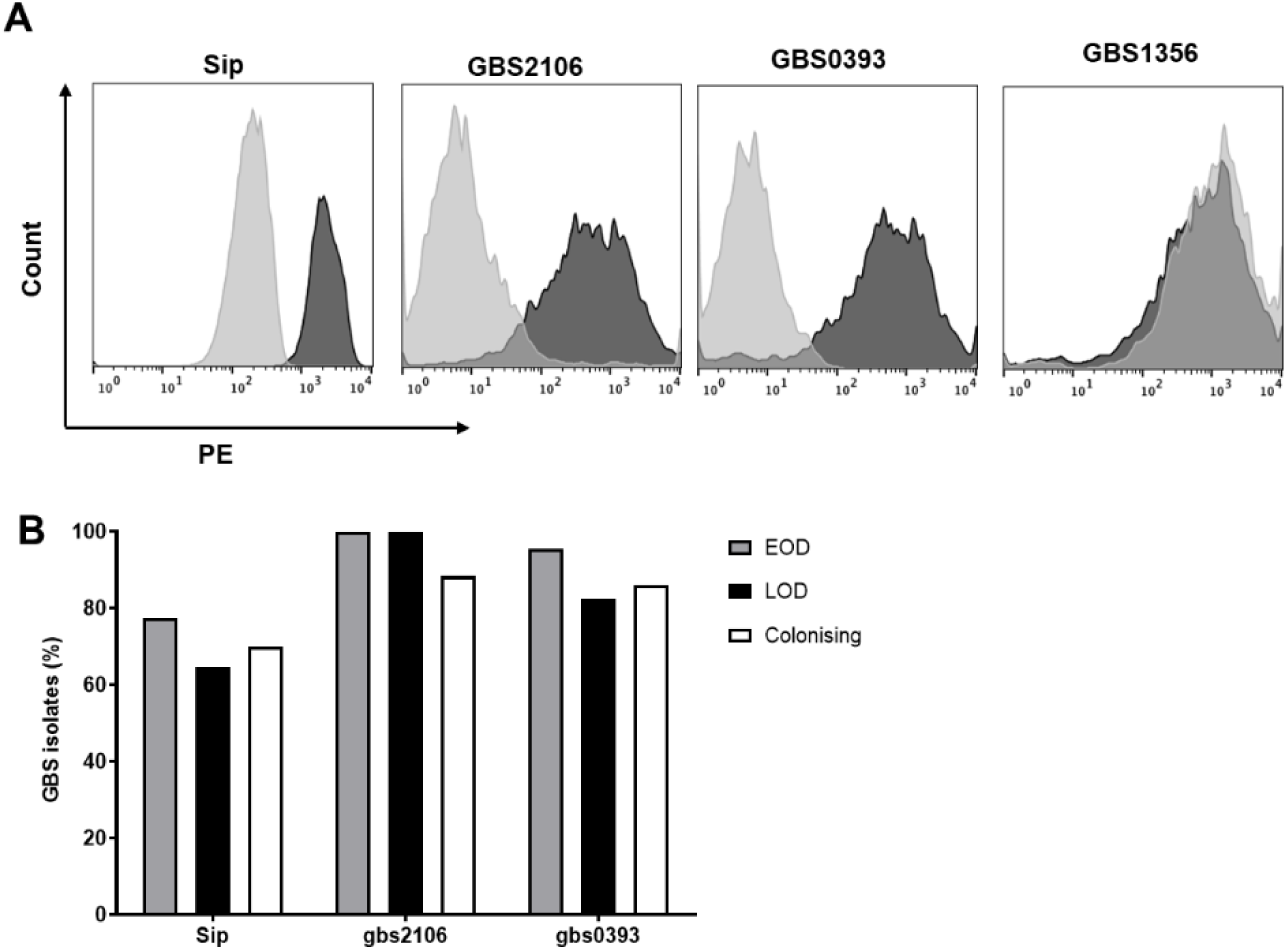
Expression of proteins on surface of Group B streptococcus (GBS) clinical isolates. A) Binding of antibodies using antigen specific antisera (red) compared to the corresponding pre-immunisation sera (blue) to access expression of each protein on the surface of GBS serotype III NEM316 strain. B) Distribution of surface proteins on GBS isolates from mothers of infants who remained healthy (white) or those that developed EOD (grey) and LOD (black).

GBS isolates collected from 83 mothers who were colonized at delivery were assessed for surface expression of proteins Sip, GBS0393 and GBS2106. Of the 83 GBS isolates screened (38 from case mothers and 43 from control mothers), Sip, GBS0393 and GBS2106 were expressed in 70.2%, 94% and 89.3% isolates, respectively. The demographic characteristics of mother-infant pairs whose colonizing GBS isolate expressed the measured proteins were similar between cases and controls (Table 1). Sip was expressed in 72.5% and 69.8% of case and controls isolates, respectively. GBS2106 protein was expressed in 100% of GBS strains isolated from mothers of cases and by 88.4% of controls. GBS0393 protein was expressed in 90.0% and 86.0% in GBS strains isolated from mothers of cases and mothers of control, respectively (Figure 1B).

### Antibody titres against GBS surface proteins in infant sera

Antibody titres against Sip, GBS0393 and GBS2106 were compared between case and control infants born to mothers colonized with GBS strains expressing homotypic proteins. IgG GMT were observed to be significantly lower in case infants compared to controls for Sip and GBS2106 (Table 2). GBS0393 specific IgG GMT were similar between the two groups of infants (Table 2). Similarly, when infants were stratified by IGbsD status and matching controls, IgG GMT for Sip and GBS2106 were lower in EOD and LOD compared to corresponding healthy controls, whereas no statistically significant differences were observed for GBS0393 IgG (Figure 2).

**Table 2:** Antibody levels against Sip, gbs2106 and gbs0393 in women of cases and controls as well as infant cases and infant controls.

|  | *protein* | *Case*<br>*(GMT, AU/mL)* | *Colonized*<br>*(GMT, AU/mL)* | *Adjusted*<br>*p-value<sup>1</sup>* | *Log-rank*<br>*p-value* |
| --- | --- | --- | --- | --- | --- |
| *infant* | sip | 64 (40,102) n=27 | 145 (102,207) n=30 | **0.008** | **0.004** |
| *infant* | gbs2106 | 61 (41,91) n=38 | 151 (107,213) n=38 | **0.001** | **0.004** |
| *infant* | gbs0393 | 119 (78,181) n=34 | 150 (100,225) n=37 | 0.928 | 0.506 |
| *mom* | sip | 212 (143,315)<br>n=27 | 268 (205,350) n=30 | 0.723 | 0.782 |
| *mom* | gbs2106 | 163 (126,210)<br>n=38 | 249 (197,313) n=38 | **0.023** | **0.084** |
| *mom* | gbs0393 | 504 (331,769)<br>n=34 | 370 (289,474) n=37 | 0.083 | 0.091 |

<sup>1</sup> adjusted for maternal age, maternal hiv status, serotype and PROM≥18h

**Figure 2.**
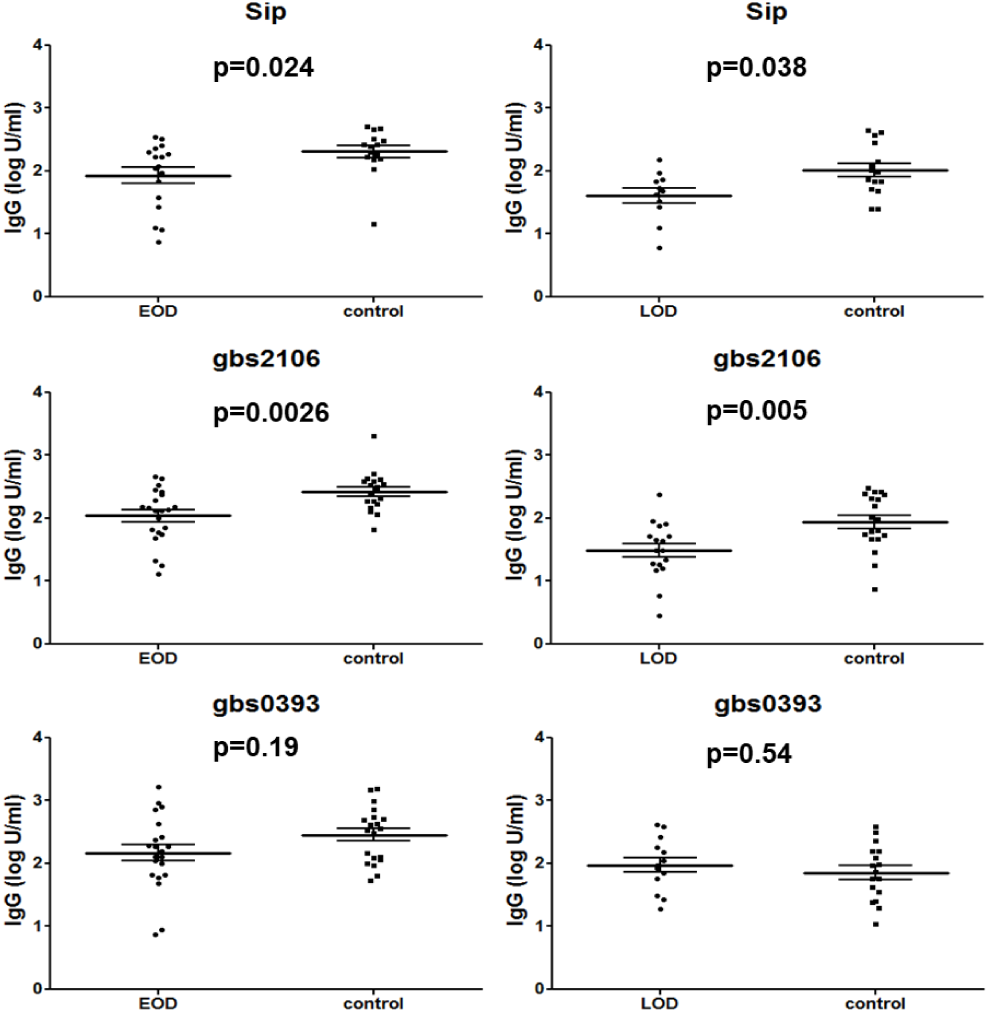
Comparison of IgG against GBS surface in infants born to mothers colonised with GBS stain expressing the respective protein between those who developed EOD or LOD and their corresponding controls.

Since the reverse cumulative plots (RCP) of antibodies were significantly different between case and control infants for Sip and GBS2106 (Figure 3A & B), we estimated absolute disease risk (ADR) for IgG titres against Sip and GBS2106 in infants using parametric and non-parametric Bayesian modelling. Figure 3C shows the parametric and non-parametric estimation of ADR associated with Sip antibodies. The parametric estimated ADR decreases with increasing Sip antibodies and steadily levels off at 2 cases per 1000 live births. Antibody titres associated with a 90% ADR reduction were not found using the parametric model. Although for non-parametric modelling, the estimated ADR continuously decreased with increasing antibody titres, and titres of 260 U/mL and 330 U/mL were associated with an 80% and 90% ADR reduction, respectively (Figure 3C). For GBS2106, the parametric estimation of the ADR monotonically decreased, and antibody titres associated with 80% and 90% reduction are 360 U/mL and 560 U/mL, respectively (Figure 3D). The non-parametric ADR curve decreases with increasing antibody titres, although as antibody titres approaches 400 U/mL, ADR increases as cases and control RCP converges. Due to the convergent of RCP for cases and controls as GBS2106 titres exceed 400 U/mL, the non-parametric estimated antibody titre associated with 80% and 90% reductions was both approximately 460 U/mL (Figure 3D).

**Figure 3.**
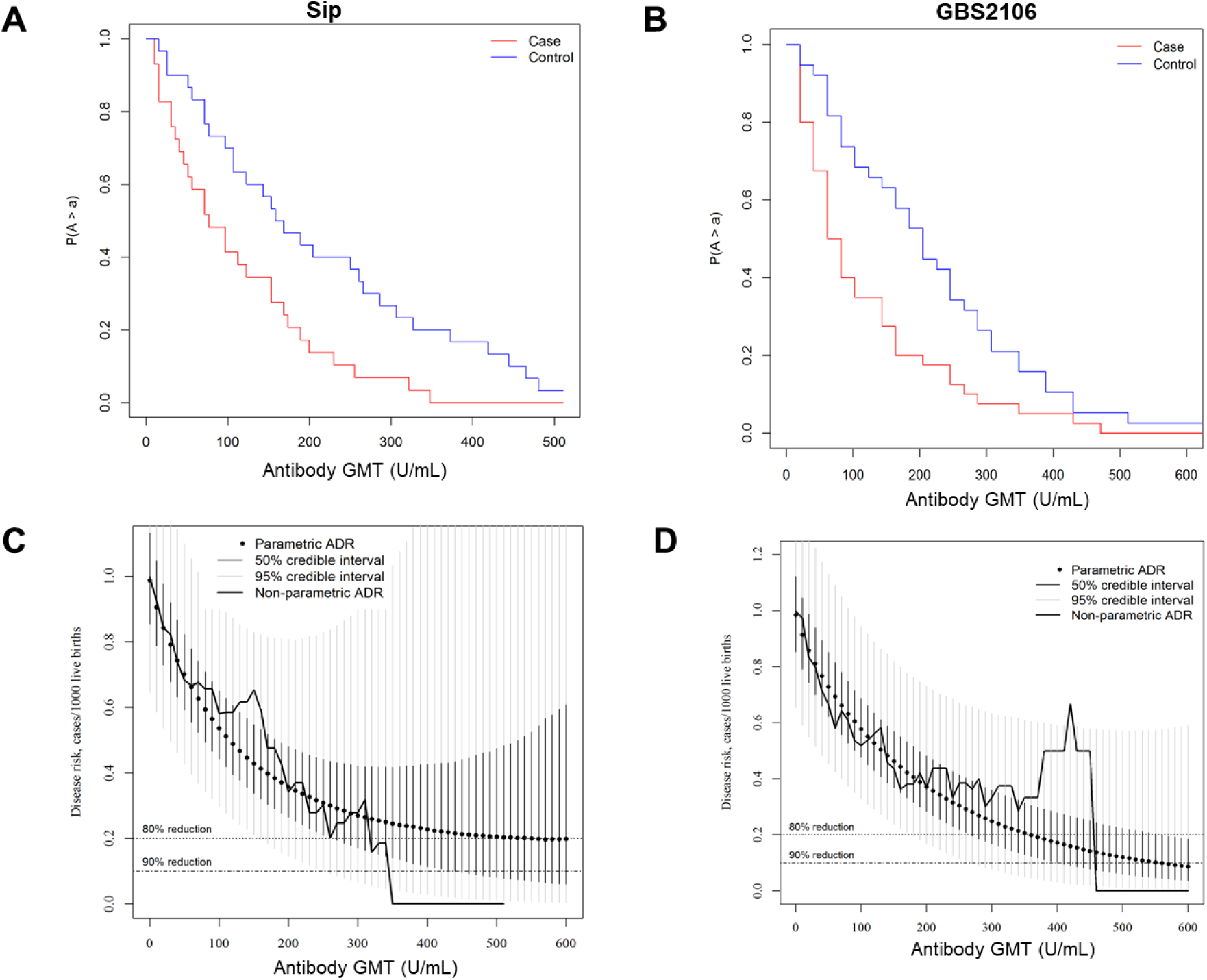
Infant IgG titre associated with reduced risk of invasive group B Streptococcus disease (IGbsD). A & B) Reverse cumulative distribution of IgG GMT (U/mL) against Sip and gbs02106 proteins in infant (<90 days of age) that remained healthy (blue) and those developed IGbsD (red). C, D) Absolute disease risk (ADR) for IgG titres against Sip and GBS2106 in infants estimated using parametric and non-parametric Bayesian modelling

### Antibody titres against GBS surface proteins in maternal sera

When comparing antibody titres for Sip, GBS0393 and GBS2106 between mothers of cases and controls colonized with GBS strains expressing homotypic protein, we observed significantly lower IgG GMT for GBS2106 in case mothers compared to control mothers, albeit not statistically significant when adjusting for co-factor variates (Table 2). IgG GMT for Sip and GBS0393 did not differ between case mothers and case controls. Further stratification of maternal IgG GMT by infant IGbsD classification, revealed significantly lower GBS2016-specific IgG GMT in mothers of LOD compared to match-controls (Figure 4). A similar trend was observed for IgG GMT against Sip, being lower in mothers of LOD compared to controls, albeit not statistically significant (Figure 4).

**Figure 4.**
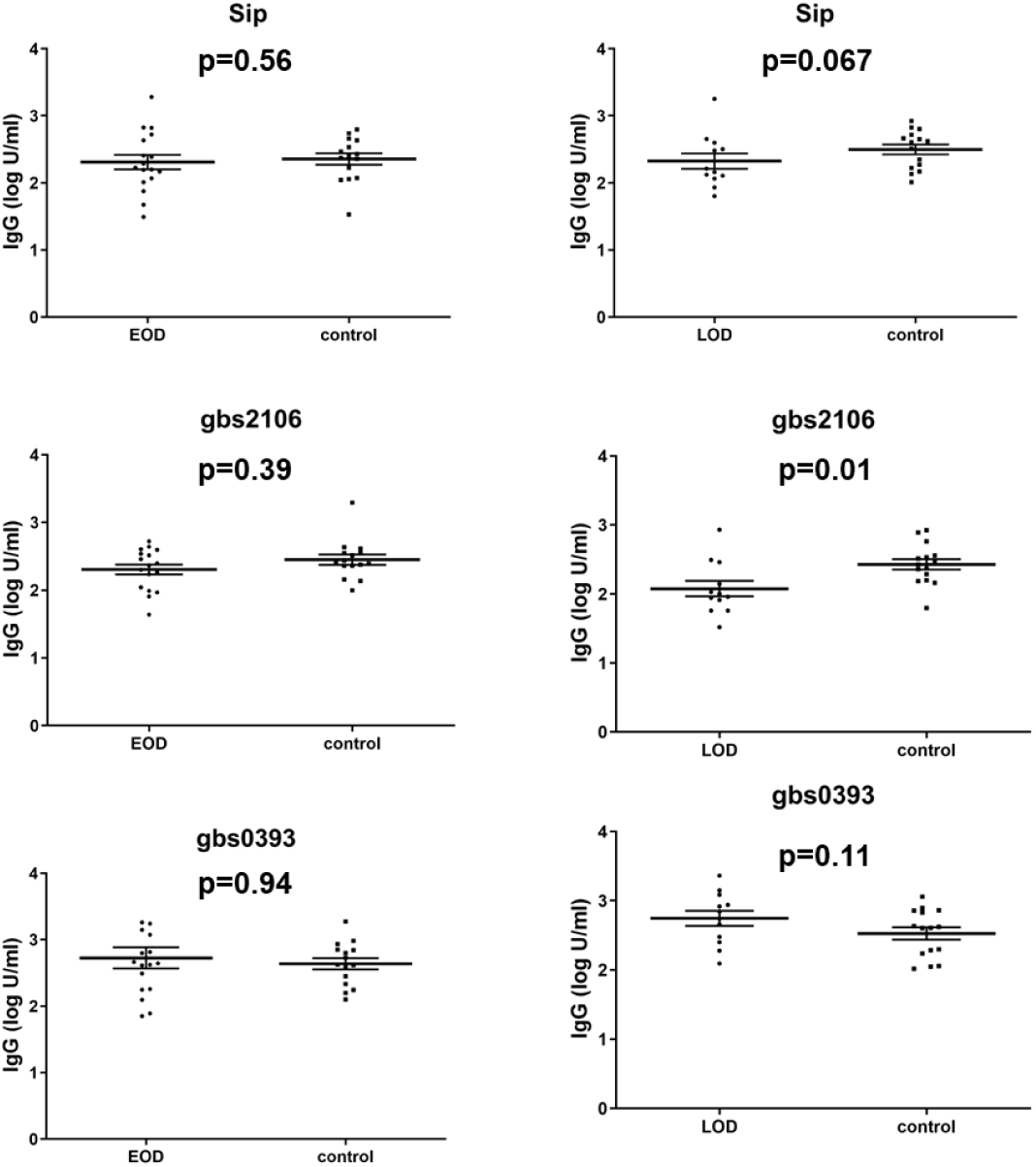
Comparison of maternal IgG against GBS surface proteins between mothers who delivered infants that developed either EOD or LOD and their corresponding controls colonized GBS strains expressing homotypic proteins.

## Discussion

Here we show that increasing antibodies for Sip and GBS2106 proteins in infants born to GBS colonised mothers were associated with reduced risk of developing invasive GBS disease. Using Bayesian models described by Carey *et al.* and Fabbrini *et al*. (18,19), putative antibody thresholds associated with 80-90% reduction of invasive GBS disease in infants were computed to be 260-330 U/mL for Sip and 360-560 U/mL for GBS2106. Although this was an explorative analysis, these findings support the role of GBS proteins contribution to disease protection corroborating murine studies (20,21). Association between naturally induced antibodies against GBS surface protein and disease risk in infants have been previously reported for Rib, FbsA and pilus island proteins (19,22,23), however no protective thresholds could be estimated in these studies.

Infants who developed EOD or LOD in turn had lower antibody titres against Sip and GBS2106 proteins compared to controls, further highlighting the importance of these proteins in GBS disease. Although Sip and GBS2016 are seemingly promising GBS vaccine candidates these results should be interpreted with extreme caution. Other factors such as gestational age, HIV infection and density of GBS colonisation could affect antibody levels in new-born infants. Trans-placental transfer of antibodies during pregnancy are highest ≥36 weeks of gestation and consequently pre-term infants would have received quantitatively lower protective antibodies. Maternal HIV infection impedes active transfer of IgG across the placenta (24) and we have previously demonstrated reduced trans-placental transfer of antibodies to GBS surface proteins including for Sip and GBS2106 of HIV-exposed uninfected infants (25). Whether colonising density of GBS in mothers increases transmission and/or risk of disease in infants remains to be known. Detection of GBS in urine of pregnant women was correlated with increasing grade of chorioamnionitis (26), a known risk factor for invasive GBS disease in their infants. Thus, we are cognisant of the possibility that the severity of GBS disease in the infants could also lead to the depletion of circulating antibodies resulting in the observed study outcomes.

In our study we used an in-house reference sera for reporting antibody titres and protective threshold that would otherwise vary by study setting and assay method. These proteins would need to be tested using standardized reference sera to determine universal protective threshold if any. Also, the observed association between protein specific antibodies and invasive GBS disease in our setting could be a proxy for capsular polysaccharide immunity previously shown to reduce the risk of GBS disease (10). Therefore, future studies would need to measure antibody-mediated killing induced by GBS proteins to further strengthen their role in protection against invasive disease.

Given the low incidence of GBS disease in the population, attempts to prove efficacy of a vaccine would require an infeasibly large sample size that would ultimately impeding vaccine licensure (27). An alternative pathway ought to expedite vaccine licensure would be to establish antibody levels associated with reduced risk of invasive GBS disease in infants. This approach has been illustrated in several studies which have reported that infants born to mothers with low circulating antibodies against GBS capsular polysaccharides have an increased risk of developing EOD GBS disease (7–10,19,28).

Therefore, in order for an antigen to be considered as a potential vaccine target against invasive GBS disease based on serological correlates of protection; the antigen should (i) be highly immunogenic, (ii) antigen-specific antibodies must efficiently cross the placenta, (iii) confer protection to both the mother and infant, (iv) antibodies must persist throughout infancy and (iv) possess a broad-spectrum target against disease causing GBS serotypes. Sip and GBS2106 are highly promising vaccine candidates, fulfilling at least 4/5 of the stipulated criteria. Both proteins were shown to be highly immunogenic GBS proteins (29), and in the absence of maternal HIV-infection, IgG against these proteins were efficiently transferred across the placental (30). Manning *et al*. reported that antibodies against Sip in infants persisted throughout infancy being detectable up until 6 months of age (31). Among the GBS clinical isolates analysed for the surface exposure of Sip and GBS2106, ≥71% isolates across all serotypes expressed these proteins. Here, we demonstrated that increasing maternal antibodies were associated with GBS disease reduction. Moreover, an oral based vaccine formulation of Sip was able to clear GBS vaginal colonization (32). We have previously shown that acquisition of GBS colonisation at the female genitourinary tract significantly increased systemic antibodies against Sip more so than when GBS acquisition occurred at the rectal tract (33). Therefore, Sip and gbs2106 fulfil most if not all criteria for an ideal GBS vaccine warranting inclusion as future vaccine candidates.

## Data Availability

All data produced in the present study are available upon reasonable request to the authors.

## Notes

### Competing Interest Statement

The authors have declared no competing interest.

### Funding Statement

This work was supported by the National Research Foundation/Department of Science and Technology: South Africa Research Chair Initiative in Vaccine Preventable Disease. The founders did not partake in the study design, data analysis, or the decision to submit for publication.

### Author Declarations

Human Research Ethics Committee at the University of Witwatersrand gave ethical approval for this work(Approval number: M140902).

## References

1. Edmond KM, Kortsalioudaki C, Scott S, Schrag SJ, Zaidi AKM, Cousens S, et al. Group B streptococcal disease in infants aged younger than 3 months: systematic review and meta-analysis. Lancet. 2012 Jan 4;379(9815):547–56.

2. Le Doare K, Heath PT. An overview of global GBS epidemiology. Vaccine. 2013 Aug 28;31 Suppl 4:D7–12.

3. Dagnew AF, Cunnington MC, Dube Q, Edwards MS, French N, Heyderman RS, et al. Variation in reported neonatal group B streptococcal disease incidence in developing countries. Clin Infect Dis. 2012;55(1):91–102.

4. Liu L, Oza S, Hogan D, Perin J, Rudan I, Lawn JE, et al. Global, regional, and national causes of child mortality in 2000–13, with projections to inform post-2015 priorities: an updated systematic analysis. Lancet. 2015;385(9966):430–40.

5. Baker CJ, Edwards MS, Kasper DL. Role of antibody to native type III polysaccharide of group B Streptococcus in infant infection. Pediatrics. 1981 Oct;68(4):544–9.

6. Baker CJ, Recnh MA, Kasper DL. Response to Type III Polysaccharide in Women whose Infants have had Invasive Group B Streptococcal Infection. N Engl J Med. 1990;322(26):1857– 60.

7. Lin F-YCY, Philips III JB, Azimi PH, Weisman LE, Clark P, Rhoads GG, et al. Level of maternal antibody required to protect neonates against early-onset disease caused by group B Streptococcus type Ia: a multicenter, seroepidemiology study. J Infect Dis. 2001 Oct 15;184(8):1022–8.

8. Lin F-YC, Weisman LE, Azimi PH, Philips JB, Clark P, Regan J, et al. Level of maternal IgG anti-group B streptococcus type III antibody correlated with protection of neonates against early-onset disease caused by this pathogen. J Infect Dis. 2004 Sep 1;190(5):928–34.

9. Baker CJ, Carey VJ, Rench M a., Edwards MS, Hillier SL, Kasper DL, et al. Maternal antibody at delivery protects neonates from early onset group B streptococcal disease. J Infect Dis. 2014;209(5):781–8.

10. Dangor Z, Kwatra G, Izu A, Adrian P, Cutland CL, Velaphi S, et al. Correlates of protection of serotype-specific capsular antibody and invasive Group B Streptococcus disease in South African infants. Vaccine. 2015;33(48):6793–9.

11. Baker CJ, Paoletti LC, Wessels MR, Guttormsen HK, Rench M a, Hickman ME, et al. Safety and immunogenicity of capsular polysaccharide-tetanus toxoid conjugate vaccines for group B streptococcal types Ia and Ib. J Infect Dis. 1999 Jan;179(1):142–50.

12. Kotloff KL, Fattom a, Basham L, Hawwari a, Harkonen S, Edelman R. Safety and immunogenicity of a tetravalent group B streptococcal polysaccharide vaccine in healthy adults. Vaccine. 1996 Apr;14(5):446–50.

13. Madhi SA, Cutland CL, Jose L, Koen A, Govender N, Wittke F, et al. Safety and immunogenicity of an investigational maternal trivalent group B streptococcus vaccine in healthy women and their infants: a randomised phase 1b/2 trial. Lancet Infect Dis. 2016;3099(16):1–12.

14. Absalon J, Segall N, Block SL, Center KJ, Scully IL, Giardina PC, et al. Safety and immunogenicity of a novel hexavalent group B streptococcus conjugate vaccine in healthy, non-pregnant adults: a phase 1/2, randomised, placebo-controlled, observer-blinded, dose-escalation trial. Lancet Infect Dis. 2021;21(2):263–74.

15. Buurman ET, Timofeyeva Y, Gu J, Kim JH, Kodali S, Liu Y, et al. A Novel Hexavalent Capsular Polysaccharide Conjugate Vaccine (GBS6) for the Prevention of Neonatal Group B Streptococcal Infections by Maternal Immunization. J Infect Dis. 2019;220(1):105–15.

16. Dzanibe S, Adrian P V, Kimaro SZ, Madhi SA. Natural acquired group B Streptococcus capsular polysaccharide and surface protein antibodies in HIV-infected and HIV-uninfected children. Vaccine. 2016;

17. Pickering JW, Martins TB, Greer RW, Schroder MC, Astill ME, Litwin CM, et al. A multiplexed fluorescent microsphere immunoassay for antibodies to pneumococcal capsular polysaccharides. Am J Clin Pathol. 2002 Apr;117(4):589–96.

18. Carey VJ, Baker CJ, Platt R. Bayesian inference on protective antibody levels using case-control data. Biometrics. 2001;57(1):135–42.

19. Fabbrini M, Rigat F, Rinaudo CD, Passalaqua I, Khacheh S, Creti R, et al. The Protective Value of Maternal Group B Streptococcus Antibodies: Quantitative and Functional Analysis of Naturally Acquired Responses to Capsular Polysaccharides and Pilus Proteins in European Maternal Sera. Clin Infect Dis. 2016;63(6):746–53.

20. Martin D, Gagnon E, Boyer M, Charland N, Brodeur BR, Al MET, et al. Protection from Group B Streptococcal Infection in Neonatal Mice by Maternal Immunization with Recombinant Sip Protein. 2002;70(9):4897–901.

21. Brodeur BR, Martine B, Charlebois I, Hamel JJ, Couture F, Rioux CRCR, et al. Identification of Group B Streptococcal Sip Protein, Which Elicits Cross-Protective Immunity. Tuomanen EI, editor. Infect Immun. 2000;68(10):5610–8.

22. Larsson C, Lindroth M, Nordin P, Stålhammar-Carlemalm M, Lindahl G, Krantz I. Association between low concentrations of antibodies to protein alpha and Rib and invasive neonatal group B streptococcal infection. Arch Dis Child Fetal Neonatal Ed. 2006 Nov;91(6):F403–8.

23. Dangor Z, Kwatra G, Izu A, Adrian P, Cutland CL, Velaphi S, et al. Association between maternal Group B Streptococcus surface-protein antibody concentrations and invasive disease in their infants. Expert Rev Vaccines. 2015;14(12):1651–60.

24. de Moraes-Pinto MI, Verhoeff F, Chimsuku L, Milligan PJ, Wesumperuma L, Broadhead RL, et al. Placental antibody transfer: influence of maternal HIV infection and placental malaria. Arch Dis Child Fetal Neonatal Ed. 1998;79(3):F202–5.

25. Dzanibe S, Adrian PV, Kimaro Mlacha SZ, Dangor Z, Kwatra G, Madhi SA. Reduced transplacental transfer of group b streptococcus surface protein antibodies in HIV-infected mother-newborn dyads. J Infect Dis. 2017;215(3).

26. Anderson BL, Simhan HN, Simons KM, Wiesenfeld HC. Untreated asymptomatic group B streptococcal bacteriuria early in pregnancy and chorioamnionitis at delivery. Am J Obstet Gynecol. 2007;196(6):524.e1–524.e5.

27. Madhi SA, Dangor Z, Heath PT, Schrag S, Izu A, Meulen AS, et al. Considerations for a phase-III trial to evaluate a group B Streptococcus polysaccharide-protein conjugate vaccine in pregnant women for the prevention of early- and late-onset invasive disease in young-infants. Vaccine. 2013;31(Supplementary 4):D52–7.

28. Matsubara K, Katayama K, Baba K, Nigami H, Harigaya H, Sugiyama M. Seroepidemiologic studies of serotype VIII group B Streptococcus in Japan. J Infect Dis. 2002;186(6):855–8.

29. Meinke AL, Senn BM, Visram Z, Henics TZ, Minh DB, Schüler W, et al. Immunological fingerprinting of group B streptococci: from circulating human antibodies to protective antigens. Vaccine. 2010 Oct 8;28(43):6997–7008.

30. Dzanibe S, Adrian P V., Kimaro Mlacha SZ, Dangor Z, Kwatra G, Madhi SA. Reduced transplacental transfer of group b streptococcus surface protein antibodies in HIV-infected mother-newborn dyads. J Infect Dis. 2017;215(3):415–9.

31. Manning SD, Wood S, Kasha K, Martin D, Rioux S, Brodeur B, et al. Naturally occurring antibodies for the group B streptococcal surface immunogenic protein (Sip) in pregnant women and newborn babies. Vaccine. 2006 Nov 17;24(47–48):6905–12.

32. Diaz-Dinamarca DA, Soto DA, Leyton YY, Altamirano-Lagos M. Oral vaccine based on a surface immunogenic protein mixed with alum promotes a decrease in Streptococcus agalactiae vaginal colonization in a mouse model. Mol Immunol. 2018;103:63–70.

33. Dzanibe S, Kwatra G, Adrian PV, Kimaro-Mlacha SZ, Cutland CL, Madhi SA. Association between antibodies against group B Streptococcus surface proteins and recto-vaginal colonisation during pregnancy. Sci Rep. 2017;7(1).

